# Using a Novel Digital Go/No-Go to Dissociate Intra-subject Temporal Fluctuations in Reaction Time and Accuracy

**DOI:** 10.1101/2024.06.12.24308834

**Authors:** Theresa M. Nguyen, Mindy K. Ross, Emma Ning, Sarah Kabir, Andrea T. Cladek, Amruta Barve, Ellyn Kennelly, Faraz Hussain, Jennifer Duffecy, Scott L. Langenecker, John Zulueta, Alexander P. Demos, Olusola A. Ajilore, Alex D. Leow

## Abstract

Impulsivity can be a risk factor for serious complications for those with mood disorders. To understand intra-individual impulsivity variability, we analyzed longitudinal data of a novel gamified digital Go/No-Go (GNG) task in a clinical sample (n=43 mood disorder participants, n=17 healthy controls) and an open-science sample (n=121, self-reported diagnoses). With repeated measurements within-subject, we disentangled two aspects of GNG: reaction time and accuracy in response inhibition (i.e., incorrect No-Go trials) with respect to diurnal and potential learning effects. Mixed-effects models showed diurnal effects in reaction time but not accuracy, with a significant effect of hour on reaction time in the clinical sample and the open-science sample. Moreover, subjects improved on their response inhibition but not reaction time. Additionally, significant interactions emerged between depression symptom severity and time-of-day in both samples, supporting that repeated administration of our GNG task can yield mood-dependent circadian rhythm-aware biomarkers of neurocognitive function.

## INTRODUCTION

For patients with psychiatric disorders, impulsivity can lead to disruptive, costly, and potentially harmful outcomes. Broadly defined and multi-faceted with various potential subtypes [1], impulsivity has been reported to be elevated in various psychiatric disorders in conjunction with altered executive functioning. Although signs of impulsivity may manifest in any individual, regardless of whether or not they carry a formal psychiatric disorder diagnosis, it is more commonly associated with select psychiatric disorders (e.g., mania in bipolar disorder [2], attention deficit hyperactivity disorder (ADHD) [3])

Although numerous studies have investigated the effects of impulsivity within specific psychiatric disorders [4–11], few studies have examined intra-individual time courses of impulsivity using a naturalistic in-situ approach (i.e., fluctuations throughout the day or ‘diurnal patterns’). Those who have examined these fluctuations primarily used ecological momentary assessments (EMAs), a subjective self-report measure, have found that increased subjective impulsivity ratings were associated with greater negative affect and suicide risk ratings, worse medication adherence, poorer cognitive functioning [12], greater impulsivity measures for those with bipolar disorder [13], and greater intra-individual variability for impulsivity for participants with bipolar disorder [14]. Given that impulsivity is not a constant state within each person, intra-individual variability in impulsivity is highly relevant, making impulsivity particularly suitable for a dimensional approach especially when viewed trans-diagnostically [1]. An additional factor to consider when examining impulsivity is trait, a constant characteristic, versus state impulsivity. Although impulsivity is a major component of manic episodes (i.e., state-dependent deficit), it can continue to persist in euthymia (trait) or other mood phases [2, 15, 16].

Current neurocognitive tasks used to measure impulsivity, and therefore executive control and response inhibition, are the Go/No-Go (GNG) task [17], the stop-signal reaction time task [18], and the Flanker Task [19]. These tasks assess different cognitive processes, and the GNG task specifically measures response inhibition [20], a central component of executive functioning [21–25]. The classic GNG task paradigm has two trials: “Go” and “No-Go” trials, where the participant is required to perform an action for “Go” trial stimuli and to omit this action for “No-Go” trial stimuli. Inhibitory control is measured by the probability of incorrectly responding on a “No-Go” trial. The GNG task allows for not only reaction time measurements for correct “Go” trials, but also false “No-Go” rates in addition to correct rejections for the “No-Go” trials and missed “Go” targets. Previous studies have shown that for the GNG task in particular, increased failure rates for the “No-Go” trials have been positively correlated to impulsivity on self-reports on the Barratt Impulsivity Scale, BIS-11 [26–30].

Researchers using neurocognitive tasks to measure impulsivity have focused on attention, reaction time, and errors. But measures such as reaction time variability and time-of-day may better reflect intra-individual patterns and adaptations of impulsivity and fluctuations in cognitive processes [31, 32]. Additionally, changes in routine, age, or other external factors (e.g., caffeine consumption [33], fatigue [34]) can modulate circadian (time-of-day) rhythms of cognitive performance [35]. [27]. The correlation between certain psychiatric disorders (e.g., bipolar disorder, schizophrenia, substance abuse disorder, depressive mood) and disrupted sleep and circadian rhythms [36–39] and evening chronotypes [40–48] has also been widely established. These variations in cognitive fluctuations and instability demonstrate the need for cognitive measurements to be taken at various times throughout the day, ideally using an in-situ naturalistic approach, especially for participants with psychiatric disorders who may have the evening chronotype.

To illustrate the importance of taking a naturalistic in-situ approach, previous studies deploying these neurocognitive tasks have primarily administered them during in-person visits, which is limited by the frequency of visits and likely restricts the hours in which these tasks can be completed and increases the risk of performance anxiety. With the increasing ubiquity of smartphones, researchers have been able to adapt neurocognitive tasks, such as the Psychomotor Vigilance Test (PVT) [49, 50], the GNG [51–53], and The Flanker Task [4, 54], to smartphones to measure individual changes in reaction times, inhibitory control, and alertness in-the-wild. To this end, a few studies in human-computer interaction (HCI) have used these digital adaptations to measure time-of-day fluctuations in cognitive processes, specifically alertness. Abdullah et al., measured alertness in-situ by adapting and implementing a digital smartphone version of PVT, a task measuring alertness and vigilance in healthy controls [49]. They determined that the average individuals’ alertness and performance varied up to 30% throughout the day and were able to formulate a predictive model for alertness states by correlating smartphone usage and alertness levels. Additionally, Dingler et al., adapted a digital, gamified version of the PVT, GNG, and Multiple Object Tracking tasks on smartphones to measure time-of-day fluctuations in alertness in healthy controls [51]. They found that reaction times increased for every subsequent hour that passed for all three tasks, and the higher a participant self-reported “Sleepiness” and, by extension, decreased alertness, the worse their performance in terms of higher reaction times. These two studies have established the possibility of monitoring time-of-day cognitive fluctuations via smartphones in-the-wild. However, they did so primarily in the context of HCI and thus did not investigate cognitive fluctuations in psychiatric patient populations, a more critical population with potentially more extreme chronotypes, nor were they able to disentangle the longitudinal time courses of reaction time and impulsivity. Another relationship possibly impacted by time-of-day is the trade-off between task speed and accuracy [55], which may be modulated by circadian fluctuations in dopamine [56], a key neurotransmitter for physical movement [57].

To this end, in this paper we report findings from a gamified GNG task, which allowed participants to compete against themselves due to immediate performance feedback, as part of a larger iPhone research study to examine the relationship between cognition and mood disorders and assess potential time-of-day correlations. Screenshots of the digital GNG task are shown in Figure 1. Additionally, this digital GNG adaptation allowed us to crowd-source sufficient data for longitudinal within-subject comparisons. As reaction time and response inhibition have previously been shown to be affected differently by temporal preparation, the voluntary orientation of attention [58], in those with high impulsivity compared to those with low impulsivity [59]. we set out to compare the longitudinal trajectory of reaction time (time to respond to a correct “Go” trial) to that of impulsivity (failure rates of “No-Go” trials, i.e., inability to inhibit inappropriate responses), in the context of not only the diurnal pattern within a day but also the potential learning effect across different days. We expect that there may exist a more complex relationship between these two concepts, with a potential decoupling between characteristics of their respective longitudinal trajectories. In particular, the human reaction time, while exhibiting diurnal patterns (fastest during mid-day), should not however exhibit a learning effect (i.e., unlikely to substantially improve over time), while by contrast we posit a possible learning effect in the failure rate of “No-Go” trials.

**Figure 1.**
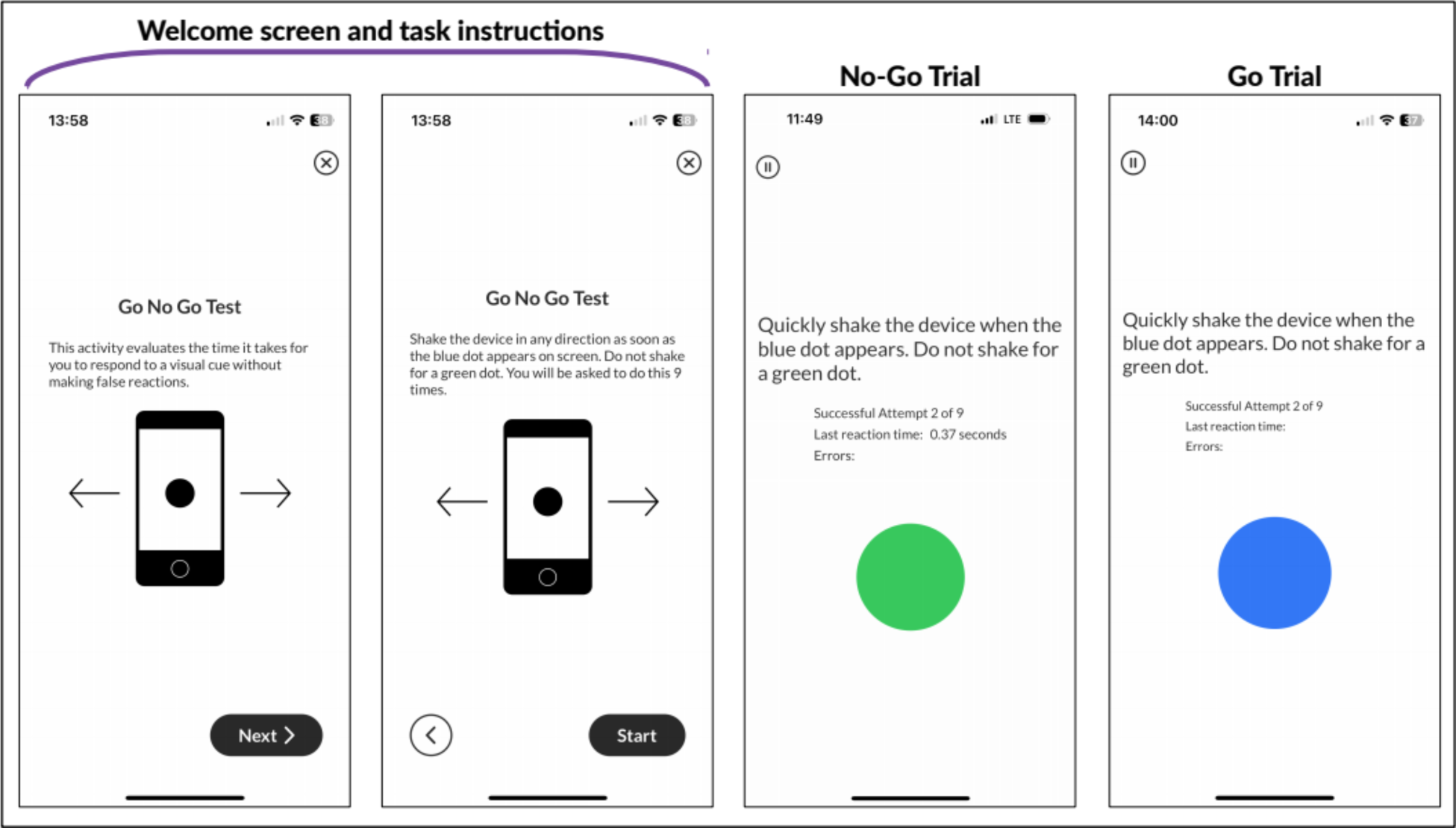
Screenshots of task introduction, instructions, and the two types of trials presented in the BiAffect go/no-go task.

## RESULTS

### Population Metrics and Mixed-effects Models for the First Sample

17 participants (mean age 30.12 ± 6.7 years) without a mood disorder and 43 participants with a mood disorder (mean age 32.6 ± 6.8), were included in the first sample. A complete overview of the demographics, diagnostic, and task contributions of the participants are listed in Table 1. This first dataset had no significant differences between the healthy control and mood disorder groups in gender, age, or number of GNG tasks completed (Table 1). There was a higher proportion of females in the mood disorder group compared to the roughly even split of males and females in the healthy control group. Participants completed approximately 25 GNG tasks on average with an adherence rate around 83%. The participants did have a significant difference in their QIDS score between the two groups (*p* < .001); those in the mood disorder group were on average mildly depressed (mean QIDS score = 5.81) compared to the healthy control group, which reported minimal depression on average (mean QIDS score = 1.06).

**Table 1.** Demographics, diagnostic, and task contributions of participants from sample one.

|  | Healthy Control Group | Mood Disorder Group | *p*-values |
| --- | --- | --- | --- |
| *n* | 17 | 43 |  |
| Gender = Female (%) | 9 (52.9) | 33 (76.7) | 0.134 |
| Age | 30.12 (6.7) | 32.64 (6.75) | 0.197 |
| QIDS Score | 1.06 (1.98) | 5.81 (4.52) | < 0.001 |
| Number of GNGs | 23.94 (6.91) | 25.86 (8.2) | 0.398 |

Note: Values are presented as mean (standard deviation) unless otherwise noted.
Abbreviations: QIDS = Quick Inventory of Depressive Symptomatology, GNG = Go/No-Go task

Mixed-effects models were constructed to predict the accuracy of “No-Go” trials (i.e. examination of commission error rates) and reaction time of correct and incorrect trial responses within the GNG tasks, summarized in Table 2. First, a logistic mixed effects model was constructed to predict the accuracy of “No-Go” trials within the GNG tasks. All of the continuous predictors were scaled, and categorical predictors were dummy-coded, so their odds ratios can be interpreted relative to one another. Odds ratios greater than 1 indicated the predictor increased the odds of a correct response on a “No-Go” trial, while odds ratios between 0 and 1 indicated the predictor decreased the odds of a correct response.

**Table 2.** Mixed effects model results for predicting “No-Go” trial accuracy and reaction time.

| Predictors | “No-Go” Trial Accuracy |  | log(Reaction Time) |  |
| --- | --- | --- | --- | --- |
|  | Odds Ratios | p-value | Log Estimate | p-value |
| Intercept | 8.399 | < .**001** | -0.717 | < .**001** |
| Age | 1.026 | 0.807 | 0.021 | 0.326 |
| Gender (Female) | 1.500 | 0.075 | 0.010 | 0.829 |
| QIDS Score | 1.041 | 0.701 | 0.037 | 0.095 |
| Learning (log(Task Number)) | 1.366 | < .**001** | 0.003 | 0.967 |
| Trail Type (“No-Go” trial) |  |  | -0.101 | < .**001** |
| Hour (1<sup>st</sup> degree) | 0.964 | 0.496 | -0.010 | < .**001** |
| Hour (2<sup>nd</sup> degree) | 1.001 | 0.526 | 0.0004 | < .**001** |
| QIDS x Hour (1<sup>st</sup> degree) | 1.046 | **0.011** | 0.005 | **0.006** |
| QIDS x Hour (2<sup>nd</sup> degree) | 1.000 | 0.840 | -0.0002 | 0.074 |
| **Random Effects (variance)** |  |  |  |  |
| Residual $\sigma^2$ | 3.290 | | 0.046 | |
| Intercept \| Subject | 0.469 |  | 0.033 |  |
| Practice \| Subject | 0.142 |  | 0.003 |  |
| **Model Fit** |  |  |  |  |
| Marginal $R^2$ / Conditional $R^2$ | 0.039/0.189 | | 0.034/0.456 | |
| log-Likelihood | -1401.653 |  | 943.100 |  |

Note: Significant p-values ( $p < .05$ ) are highlighted in bold.

As participants completed the GNG task approximately once a day, we also observed a significant effect of learning on the accuracy of their responses to “No-Go” trials (*p* < .001). Participants were 1.366 times more likely to respond more accurately on “No-Go” trials following each additional GNG task that they completed, shown in 2A. The improvement began rapidly during the first 10 tasks before beginning to plateau following a logarithmic trend as more tasks were completed. We next investigated the effect of time of day and depression severity on participants’ response accuracy to “No-Go” trials. Although there were no significant effects of depression severity nor hour of the day on “No-Go” trial accuracy, there was a significant interaction between depression severity and the linear effect of hour (*OR* = 1.046, *p* = 0.011). Those who were more depressed (QIDS score = 10) performed most poorly on the “No-Go” trials earliest in the day compared to those who were mildly and not depressed and increased in their accuracy as the day progressed.

On the other hand, those who were not depressed (QIDS score = 0) tended to perform more accurately earlier in the day and worse as the day progressed (Figure 2B).

**Figure 2.**
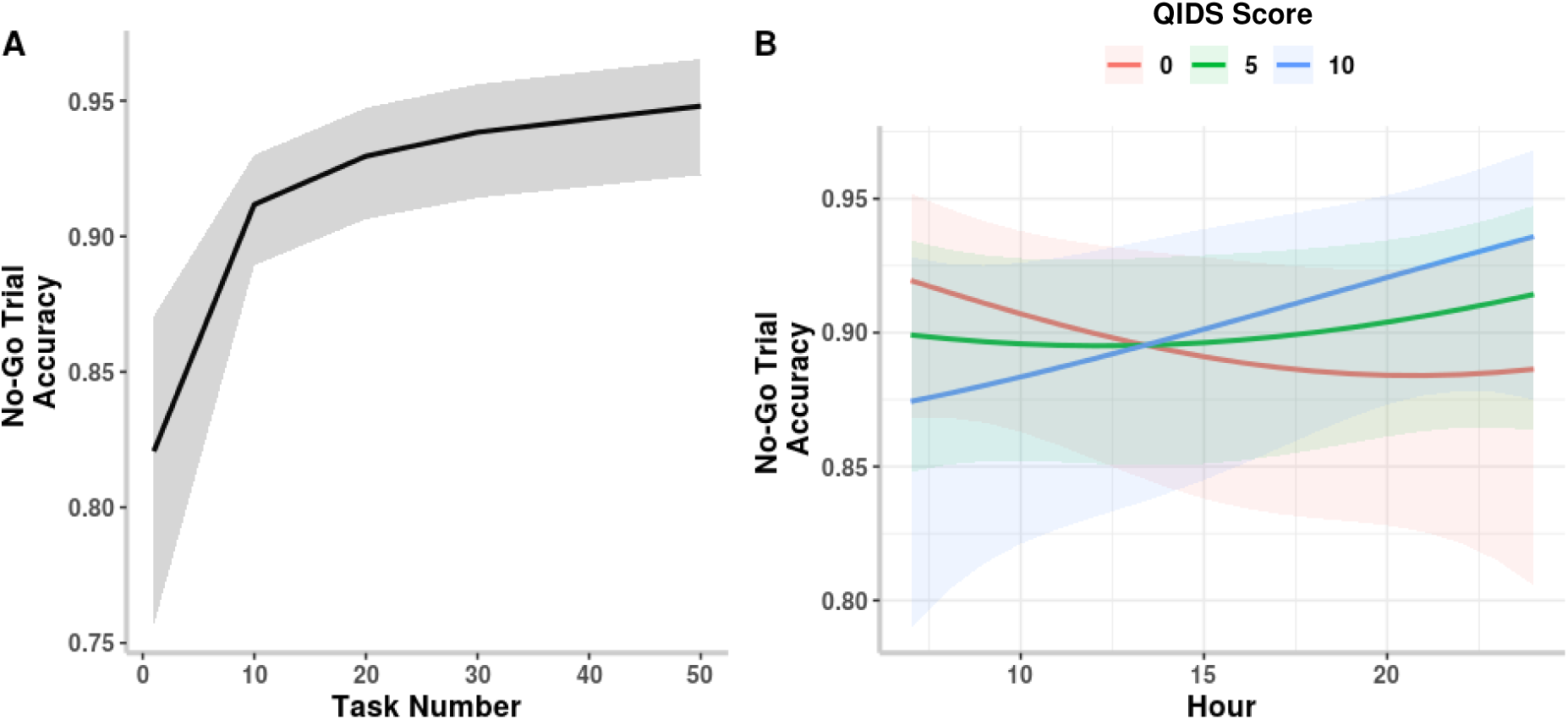
Plots showing the significant (A) effect of learning and (B) interaction between hour of the day and depression severity on “No-Go” trial accuracy.

A mixed effects model was constructed to predict the reaction time of correct “Go” and incorrect “No-Go” trials within the GNG tasks (Table 2). As with the previous model, categorical variables were dummy-coded and continuous variables were scaled, meaning that the model estimates can be interpreted as relative effect sizes. Positive slopes indicated slower reaction times, while negative slopes indicated faster reaction times. Since reaction time was log-transformed, model estimates are reported as logged estimates.

This model had a significant effect of trial type on reaction time (*B* = -0.101, *p* < .001): in the presence of incorrectly-responded “No-Go” trials, participants did so more quickly by 78 ms on average than to “Go” trials (Figure 3A), thus supporting that the BiAffect “No-Go” trials probe impulsivity. Notably, contrary to the learning effect we found in the trial accuracy analysis, there was no significant effect of learning on reaction time (*B* =0.003, *p* =0.967), meaning that subsequent GNG tasks did not result in overall faster reaction times.

**Figure 3.**
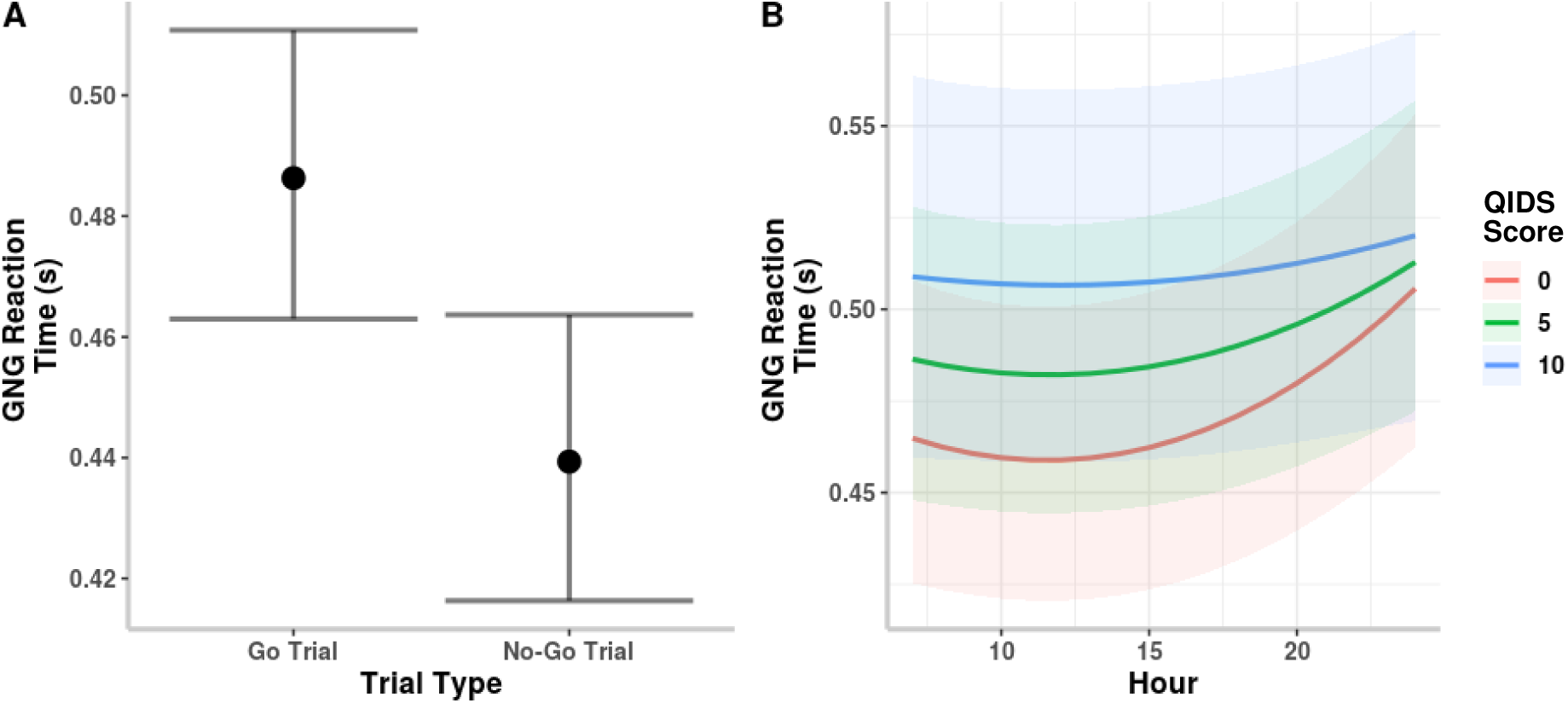
Plots showing the significant (A) effect of trial type and (B) interaction between depression severity and hour of the day on GNG reaction time.

There was a significant effect of hour (both linear and quadratic terms) on reaction time (*B* = - 0.010, *p* < .001; *B* = 0.0004, *p* < .001), suggesting that time of day quadratically affected reaction time (by contrast, we did not find a time-of-day effect in the trial accuracy analysis). Consistent with a diurnal pattern, participants responded more quickly during the middle of the day compared to the very beginning and end of the day. Moreover, although depression severity itself was not predictive of reaction time, the level of depression severity affected how time of day impacted participants’ reaction times. There was a significant interaction between depression severity and hour of the day (*B* = 0.005, *p* = 0.006). Participants who experienced no depression (QIDS score = 0) saw more well-defined diurnal dependence of their reaction times on the time of day than participants experiencing higher levels of depression (attenuated diurnal dependence). In particular, participants who were moderately depressed (QIDS score = 10) had little change in their reaction times depending on the time of day, shown in Figure 3B.

### Complementary Analyses in the Second Sample

121 participants (mean 42.21 ± 14.02 years, 67.8% female) voluntary joined the study and self-reported their diagnoses for our second sample. An additional analysis was performed to evaluate the extent of the task’s sensitivity to time of day on a more age-diverse sample. A larger sample for analysis was obtained in the second sample as described in Table 3. These participants had a similar split in gender with slightly more females than males in the sample. The mean age in this sample was about 10 years older than the previous sample, and over half of participants reported a diagnosis of a mood disorder as well as other co-morbid diagnoses.

**Table 3.** Demographic, diagnostic, and data contribution information from the open-science participants.

|  |  |
| --- | --- |
| n | 121 |
| Gender (Female (%)) | 82 (67.8) |
| Age (mean (SD)) | 42.21 (14.02) |
| Bipolar Disorder (n (%)) | 64 (52.9) |
| Depressive Disorder (n (%)) | 78 (64.5) |
| Seasonal Affective Disorder (n (%)) | 13 (10.7) |
| Anxiety (n (%)) | 74 (61.2) |
| ADHD (n (%)) | 31 (25.6) |
| PTSD (n (%)) | 30 (24.8) |
| Number GNG (mean (SD)) | 11.31 (8.36) |

Note: Diagnoses were obtained via self-report with each disorder listed as a binary response, resulting in the possibility of selecting mutually exclusive diagnoses for some participants.

The models were modified slightly to account for the differences in types of data collected and greater sparsity of the collected data. In the models below, individual diagnoses were used in place of QIDS score. Predictors were also scaled or dummy-coded to increase interpretability, and all odds ratios and estimates can be interpreted as above. Overall, the findings were similar to the original models constructed to predict “No-Go” trial accuracy and reaction time.

As before, we constructed a logistic mixed effects model to predict participants’ “No-Go” trial accuracy, with all results listed in Table 4. Similar to the learning effect in trial accuracy in the first sample, here participants again greatly improved their “No-Go” trial accuracy following subsequent completions of the GNG task as shown by the significant effect of learning in the model (*OR* = 1.403, *p* < .001; Figure 4A). Additionally, having a self-reported diagnosis of a depressive disorder significantly affected “No-Go” trial accuracy (*OR* = 0.627, *p* = 0.030), such that those with a self-reported history of depression were less accurate on “No-Go” trials than participants without the disorder (Figure 4B).

**Figure 4.**
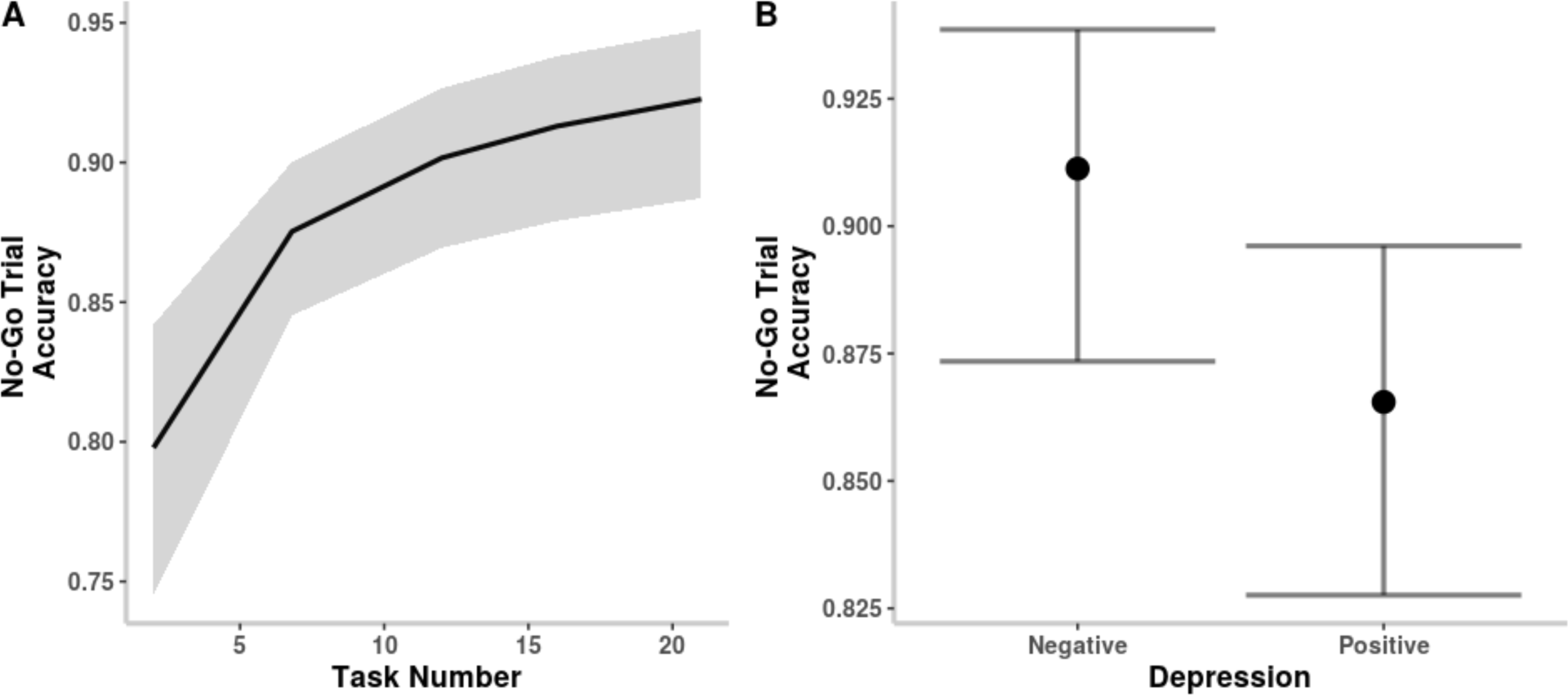
Plots of the significant main effects of (A) practice and (B) a diagnosis of a depressive disorder on “No-Go” trial accuracy.

**Table 4.** Model results for predicting “No-Go” trial accuracy and reaction time in the second sample.

|  | “No-Go” Trial Accuracy |  | log(Reaction Time) |  |
| --- | --- | --- | --- | --- |
| Predictors | Odds Ratios | p-value | Log Estimate | p-value |
| Intercept | 8.866 | < .001 | -0.757 | < .001 |
| Age | 0.906 | 0.322 | 0.034 | **0.044** |
| Gender (Female) | 0.865 | 0.495 | 0.068 | 0.050 |
| Learning (log(Task Number)) | 1.403 | < .001 | 0.005 | 0.556 |
| Bipolar Disorder | 0.982 | 0.926 | 0.015 | 0.660 |
| Depressive Disorder | 0.627 | **0.030** | -0.025 | 0.482 |
| Seasonal Affective Disorder | 1.044 | 0.889 | 0.011 | 0.822 |
| Anxiety Disorder | 1.066 | 0.765 | -0.006 | 0.822 |
| ADHD | 1.534 | 0.063 | 0.034 | 0.357 |
| PTSD | 1.099 | 0.671 | 0.073 | 0.051 |
| Trail Type (“No-Go” trial) |  |  | -0.090 | < .001 |
| Hour (1<sup>st</sup> degree) | 1.022 | 0.219 | -0.020 | 0.069 |
| Hour (2<sup>nd</sup> degree) | 1.000 | 0.960 | 0.001 | < .001 |
| **Random Effects (variance)** |  |  |  |  |
| Residual $\sigma^2$ | 3.290 | | 0.050 | |
| Intercept \| Subject | 0.426 |  | 0.024 |  |
| Practice \| Subject | 0.127 |  | 0.004 |  |
| **Model Fit** |  |  |  |  |
| Marginal $R^2$ / Conditional $R^2$ | 0.045/0.183 | | 0.045/0.390 | |
| log-Likelihood | -1178.680 |  | 223.978 |  |

Note: Significant p-values ( $p < .05$ ) are highlighted in bold.

We additionally used this second sample which incorporated a wider age range to construct a mixed effects model predicting reaction time. With this wider range, we observed a significant effect of age on reaction time (*B* = 0.034, *p* = 0.044). Older participants had slower reaction times than their younger counterparts (Figure 5A). The other significant findings aligned with our previously constructed model predicting reaction time. As before, there was a significant effect of trial type on reaction time (*B* = -0.090, *p* < .001) in that participants responded more quickly to incorrect “No-Go” trials than to correct “Go” trials, shown in Figure 5B. We once again did not note a significant effect of learning on reaction time (*B* = 0.005, *p* = 0.556). Lastly, there was a significant quadratic effect of hour on reaction time (*B* = 0.001, *p* = < .001) with participants responding more quickly during the middle of the day than the mornings or evenings (Figure 5C).

**Figure 5.**
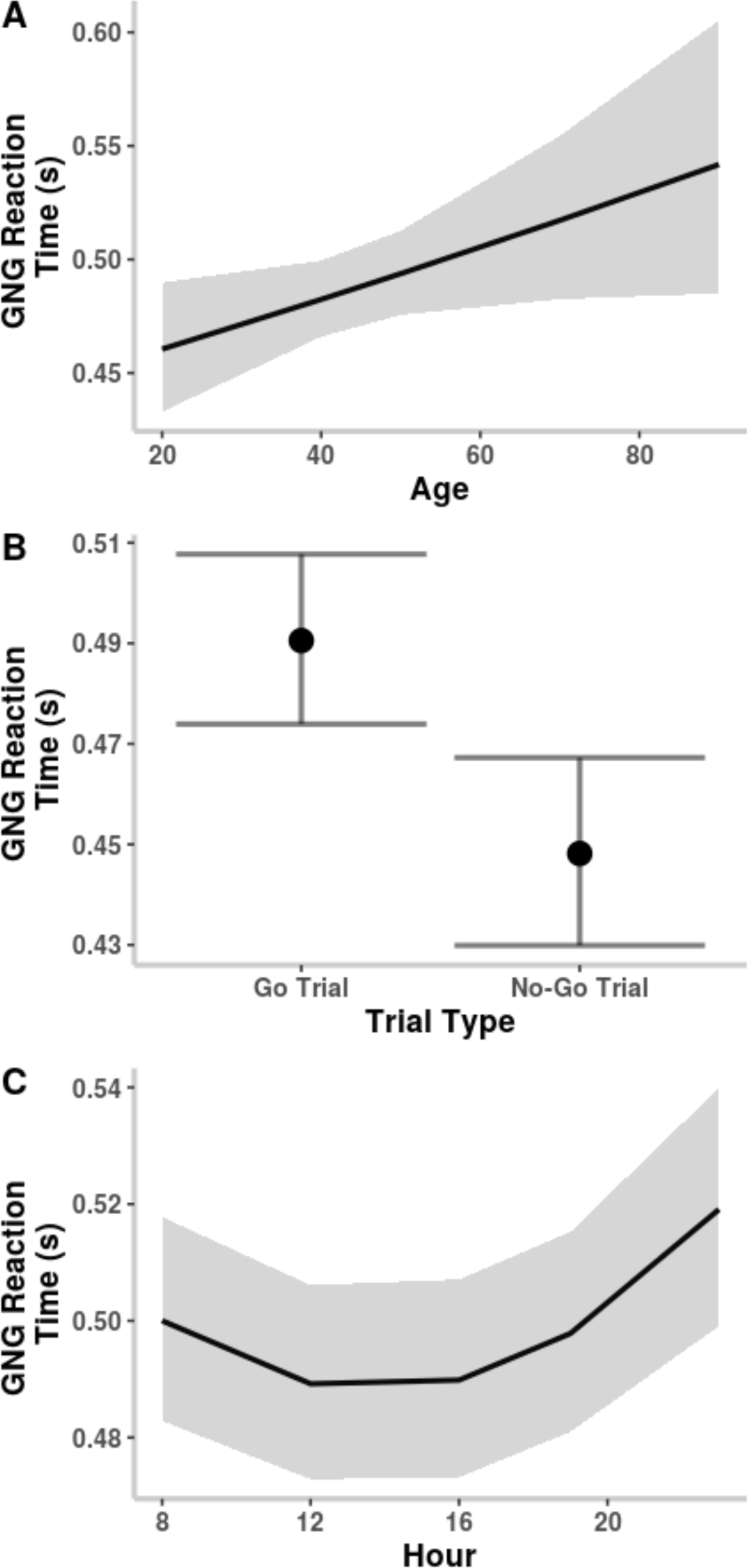
Plots showing the significant effects of (A) age, (B) trial type, and (C) hour on GNG reaction time.

## DISCUSSION

In this study, we analyzed the task performance of a short, gamified iPhone GNG task to characterize and compare the longitudinal intra-individual trajectories of reaction time and commission error rates (indexing failure to inhibit prepotent responses as an important aspect of impulsivity). By using repeated measurements within-subjects in two different samples, we were able to decouple reaction time and error rate (or alternatively accuracy) in terms of the time-of-day diurnal and any learning effects. These two samples differ in complementary ways, the first being a well-characterized but younger clinical sample, and the second innovatively utilizing crowd sourced “citizen-science” data across a wider age range and self-reported diagnoses.

First, despite the overall different strategy of recording responses (shaking the phone in “Go” trials instead of pressing a button in traditional GNG tasks) we confirmed that this short and gamified GNG task, with immediate results and improvement metrics, probes response inhibition and impulsivity, as the trial type influenced reaction time (i.e., the time it takes to reach motor threshold as recorded by accelerometry) where “Go” trials had longer times to motor threshold compared to incorrectly responded to “No-Go” trials (Figures 3A and 5B). Second, as we initially hypothesized, in both samples’ reaction time exhibited diurnal patterns (fastest during mid-day; Figures 3B and 5C) but not a learning effect (i.e., reaction time did not improve over time with additional tasks; Tables 2 and 3), while in the second sample there was also a significant age effect (slower reaction time with older age; Figure 5A). By contrast, a different pattern emerged for commission error rates in that there was a strong learning effect in both samples (subjects learned to decrease their error rates, or became more accurate and less “impulsive”, in performing this GNG task; Figures 2A and 4A), while overall we did not find a strong diurnal pattern nor an age affect (older subjects are equally accurate).

However, our results in the first sample did yield a significant time-of-day variation on GNG task accuracy dependent on the severity of depressive symptoms, shown in Figure 2B. Those with more severe depressive symptoms performed less accurately in the morning, whereas the reverse was found in those without depressive symptoms, in which accuracy was highest in the morning and decreased gradually as the day went on. In the second sample where we relied on self-reported diagnoses (but not current symptom severity), we instead found that overall people who reported a history of depression were less accurate regardless of the time of day (Figure 4B).

Additionally, in the first sample we further found a significant time-of-day first-order interaction with reaction time and depression severity (Figure 3B). In particular, those with more symptoms of depression (mild-moderate severity with QIDS of 10) had a trend towards attenuated diurnal pattern and numerically slower reaction time compared to those with QIDS of 0 (no depressive symptoms). Noting the dependence of reaction time on depression severity, we however refrain from extrapolating the data to severe depression due to mood disorder subjects in this sample were at most moderately depressed.

Many of our findings merit further discussions. First, given the age effect in reaction time (the younger the subject, the faster their reaction time) but not in accuracy, one can infer that although we may have to accept slower reaction times as we age, there is evidence that levels of performance accuracy can however be maintained (at least to the extent that is measurable using our specific GNG task). Somewhat surprisingly, we found a time-of-day main effect with respect to reaction time but not GNG accuracy, contrary to prior literature that found diurnal patterns in both [60]. A plausible explanation for a lack of main effect in our GNG accuracy is perhaps due to participants’ learning the response-inhibition aspect of the task (i.e., learning to be more careful) over time, thus attenuating any possible diurnal effect if the task were to be only administered once. Given this finding, we can more broadly postulate that certain aspects of impulsivity, independent of reaction time, could likely be learned over time. That is to say, for those with weaker response inhibition, they could have the capacity to improve their response inhibition with proper and deliberate training. Of note, this viewpoint is consistent with recent developments in digital therapeutics that aim to improve aspects of executive function through gamified cognitive training delivered via portable devices [61, 62].

Our findings of different time-of-day effects in people with depressive symptoms are also in line with current literature on chronotypes in mood disorders. In particular, the late or evening chronotype is well-recognized to be associated with more severe depressive symptoms, likely explaining why people with more severe depressive symptoms perform our GNG task less accurately in the morning [40, 43–48]. Next, despite the fact that we did not find any significant sex differences for the GNG accuracy or reaction times, some prior studies have shown that females have faster processing speed on various Woodcock-Johnson cognitive and achievement batteries [63], that females are more proactive whereas males are more reactive when responding to visuomotor tasks [64], and that females have been shown to be quicker in different speeded tasks than males [65]. It is unclear why our study did not find any sex differences, but it should be noted that our study was never specifically designed to study sex-differences.

In conclusion, this present study used a short, gamified iPhone GNG task to characterize the longitudinal trajectory of reaction time (during “Go” trials) and inhibitory control (during “No-Go” trials) using two different yet complementary samples, which when combined allowed us to explore task performance across a range of clinical and demographic variables. Our findings provided a more nuanced view of the classic speed-accuracy tradeoff. In particular, rather than a straightforward conclusion that faster reaction led to lower accuracy, we showed that with repeated task administration participants did learn to become more accurate even though they did not become faster. Also, there was a consistent time-of-day effect in reaction time but not accuracy. Moreover, while older participants were indeed slower, they were no less accurate compared to younger participants. Taken together, these findings support the idea that the neurocircuitry subserving reaction time and accuracy are likely interacting yet separate neurocognitive systems [66]. Last but certainly not least, GNG task performance appeared to be modulated by a complex interplay between depressive symptoms and the time-of-day, supporting the possibility that this gamified GNG may yield state biomarkers of neurocognitive function even with repeated task administration.

## METHODS

### UnMASCK

#### First Sample: Participants from the UnMASCK study

Participants with and without a mood disorder (healthy control = 17, 52.9% female; mood disorder = 43, 76.7% female) were recruited as part of the UnMASCK study (ClinicalTrials.gov Identifier: NCT04358900). Individuals owning iPhones between the ages of 25 and 50 were eligible for this study if they were not experiencing severe cognitive impairment, major medical or neurological illness interfering with protocol adherence, active alcohol or substance use disorders, acute suicidal ideation, or contraindications to MRI. Informed consent was obtained from all participants prior to study screening. Participants were categorized in the mood disorder group if they met the DSM-5 criteria for bipolar disorder type I/II, major depressive disorder, persistent depressive disorder, or cyclothymia as determined by a study clinician.

#### Data collection

For this analysis, participants were asked to download the BiAffect iOS app, fully developed in-house by our team, released under the official Apple Developer account of the University of Illinois, and made freely available. Once BiAffect was downloaded and properly set-up, which included an additional on-device consenting process as part of the study protocol, all participants were prompted daily to complete the BiAffect’s smartphone-based implementation of a GNG task (Figure 1).

In this gamified implementation adapted from the Reaction Time task in Apple’s original Researchkit framework (https://researchkit.org), each GNG task consists of a series of trials (in which the subject has to execute a “Go” or “No-Go” response based on a visual stimulus in the form of a colored dot, presented on screen at randomly generated time after the start of the trial), with a 3-second rest between consecutive trials. For “Go” trials, the participants were asked to shake their phone in any direction as soon as a blue dot had been presented on-screen, with the reaction time defined as the time lapse between the appearance of the blue dot and the time when the accelerometer magnitude reached the same threshold as that in the original ResearchKit Reaction Time task. For “No-Go” trials, the participants were asked to refrain from shaking their phone (inhibiting the prepotent shake response) if the color of the dot that appeared was green instead of blue. At the start of each trial, the color of the dot is randomly assigned to either green or blue with a green dot having a 1/3 probability of appearing. Throughout the entire GNG task, tri-axial accelerometry is sampled at 100 Hz, and a task terminates when the participant has achieved a total of 9 correct trials (either a blue dot responded to with a shake or a green dot not responded to). The timestamp of each trial and task, the accuracy of the trial, and the amount of time between the beginning of the trial and the response (if any) were recorded.

As part of the design of the study was to understand the relationship between diurnal patterns and GNG performance, participants who had taken the GNG task at least 10 times during their month in the study and at different hours over an 8-hour span (across all days) were included in the analysis. This restriction on the inclusion of participant data was set in order to incorporate individual differences regarding how time of day affects performance in the analysis. The severity of depression symptoms, if any, was assessed by a clinician using the Quick Inventory of Depressive Symptomology (QIDS) at the time each participant began the study as part of the overall study protocol.

#### No-Go Trial Accuracy Analysis

To analyze the effect of time of day and depression severity on “No-Go” trial accuracy within BiAffect’s GNG tasks, a logistic mixed effects model was constructed, controlling for the fixed effects of age and gender, as well as for a potential practice effect. Time of day was measured using the hour of the day in which the GNG task was completed and was truncated to include only the hours between 0700-0059 during which the majority of participants completed GNG tasks. Depression severity was measured using the participant’s QIDS score taken at study entry into the respective month-long study. A learning effect was controlled for by sequentially numbering the GNG tasks completed by each participant. These GNG task numbers were log-transformed, since we believed that participants would rapidly improve before plateauing in their performance as shown with other digital tasks [67]. “No-Go” trials were dummy-coded to be either correct (1) or incorrect (0). Gender was dummy-coded as male (0) or female (1). All continuous variables were z-scored. The random effects in the model included a random intercept per subject and random slope of learning for each participant to allow for individual differences.

#### Reaction Time Analysis

To fully characterize the BiAffect GNG task, reaction times for both correct “Go” trials and incorrect “No-Go” trials were included in this analysis. With literature supporting that human reaction time falls within the range of 220-384 ms depending on the complexity of the stimulus [68], BiAffect reaction times were filtered to only include trials with times between 200-1000 ms (0.2-1.0 s; Figure 6), thus removing accidental shakes (those shorter than 200 ms) and distraction or attentional lapse (longer than 1000 ms). The remaining times were then log-transformed to form a more normal distribution for analysis. We used a mixed effects model to determine the effect of time of day, trial type, and depression severity on reaction time, controlling for age, gender, and learning. Variables used in the “No-Go” trial accuracy model were treated the same. Trial type was dummy-coded as either a “Go” trial (0) or a “No-Go” trial (1). The random effects were the same as the previous model: random intercept per subject and random slope of learning per participant.

**Figure 6.**
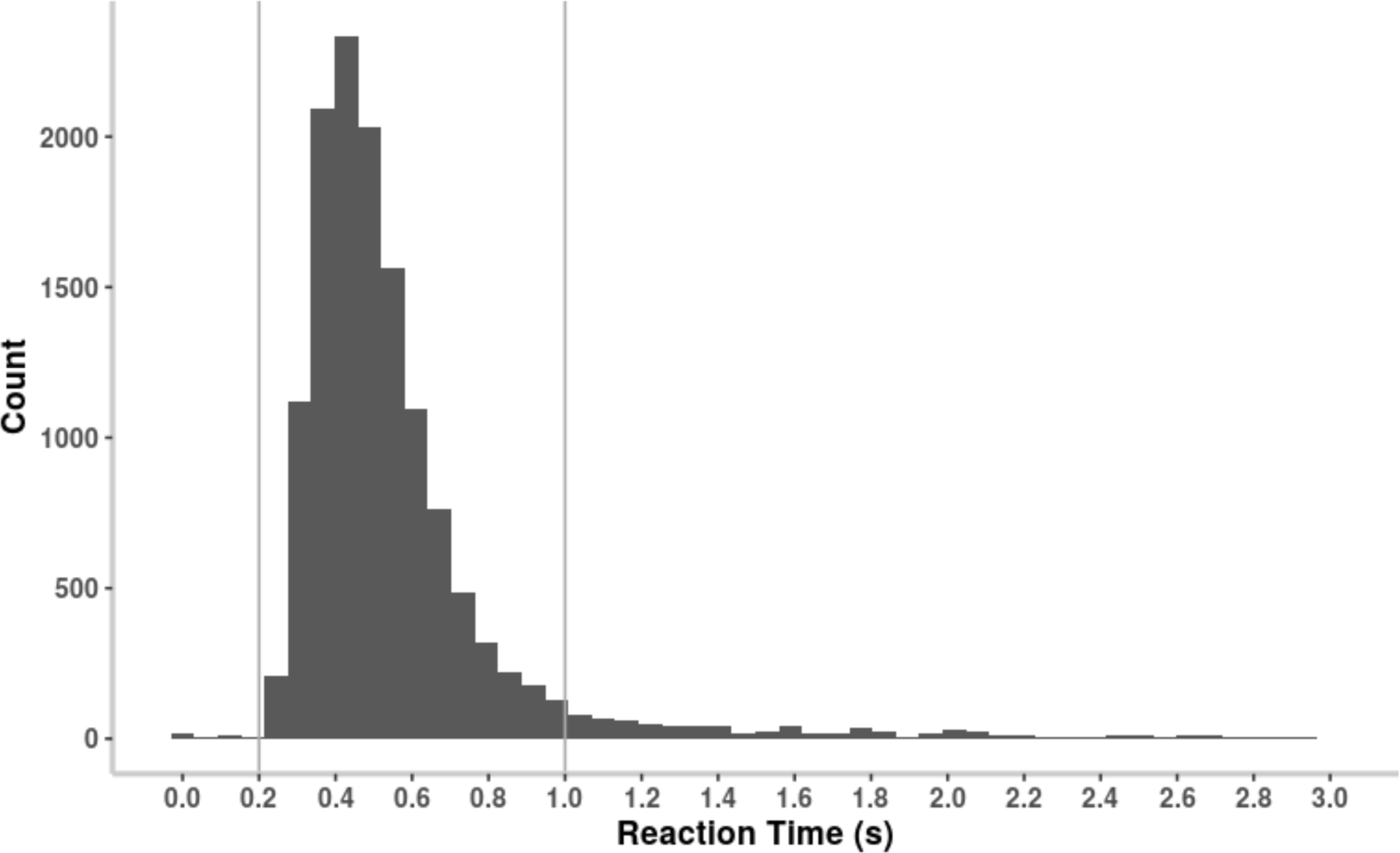
Histogram of task reaction times, across all study participants, with the filtered 0.2-1 s range marked with vertical lines.

### COMPLEMENTARY ANALYSIS IN A SECOND SAMPLE

#### Second sample: crowd-sourcing GNG data from citizen science participants

To complement our first sample, where a restricted age range meant it was more difficult to examine for a potential age effect and its interaction with other variables, we expanded our analysis to examine task performance in a larger open-science dataset of participants by crowd-sourcing BiAffect GNG task data from any individual (i.e. citizen scientist) who downloaded the BiAffect app onto their personal iPhones and gave informed consent for data collection through the app independently of the first sample described above. Due to the greater sparsity in the data, only participants who had completed the GNG task (Figure 1) at least 4 times during any hour of the day were included in these analyses. These participants consisted of those who self-reported a known diagnosis of a mental illness or did not have any known diagnoses. A total of 121 participants were included in this second sample, ranging in age from 18 to 89 years.

#### Second sample: Statistical Analyses

Since the participants in the second sample were not guided through their first GNG task, the first task from each participant in this sample was treated as a “practice run” task and removed from the analysis. The total number of tasks each participant was allowed to contribute was capped at 20 tasks, excluding the first practice task, as the median number of tasks across the entire second sample was 11. We then repeated both mixed effects models, as with the first sample, to predict “No-Go” trial accuracy and reaction time with some differences in the fixed effects compared to the previous models given the differences in data collected. The citizen science participants did not have a clinician-rated QIDS score, so each individual’s self-reported diagnoses were entered and controlled for in the model in its place. We also restricted the range in hours in the model to be between 0800-2359 to only include the hours of the day in which the majority of participants completed GNG tasks. All other variables were z-scored or dummy-coded as described above. Both models were constructed as described above in terms of their random effects.

Data processing was done in Python 3.7 [69] using the Pandas (version 1.3.5) [70, 71] and Numpy (version 1.19.2) [72] packages. All analyses were conducted in R (version 4.3.1) [73] using the tidyverse package (version 2.0.0) [74] for data processing, the lmerTest package (version 3.1-3) [75] for analysis, and the effects (version 4.2-2) [76] and ggpubr (version 0.6.0) [77] packages for visualization.

## Data Availability

All data will be available on the NIMH Data Archive.

## DATA AVAILABILITY

All data will be available on the NIMH Data Archive.

## ACKNOLWEDGEMENTS

This research was funded by the National Institute of Mental Health of the National Institutes of Health (NIMH-NIH), R01MH120186.

## AUTHOR CONTRIBUTION

T.M.N., M.K.R., E.N., F.H., J.D., S.L.L., J.Z., A.P.D., O.A.A., A.D.L. conceptualized the study. Formal analysis was done by T.M.N., M.K.R., E.N., A.P.D., A.D.L. Data acquisition was done by S.K., A.T.K., A.B., and E.K. Funding was acquired by O.A.A. and A.D.L. Supervision was provided by A.P.D., O.A.A., and A.D.L. T.M.N., M.K.R, and A.D.L. drafted the initial manuscript. All authors, T.M.N., M.K.R., E.N., S.K., A.T.C., A.B., E.K., F.H., J.D., S.L.L., J.Z., A.P.D., O.A.A., and A.D.L. were involved in reviewing and editing.

